# Gamma connectivity predicts response to intermittent Theta Burst Stimulation in Alzheimer’s disease: A randomised controlled trial

**DOI:** 10.1101/2022.02.21.22271264

**Authors:** Kate E. Hoy, Melanie R. L. Emonson, Neil W. Bailey, Caitlyn Rogers, Hannah Coyle, Freya Stockman, Paul B. Fitzgerald

## Abstract

Alzheimer’s disease is characterised by progressive cognitive decline for which there are currently no effective treatments. There is growing evidence that neural network dysfunction is a likely proximate cause of cognitive impairment in Alzheimer’s and, as such, may represent a promising therapeutic target. Here we investigated whether a course of intermittent Theta Burst Stimulation (iTBS) could modulate functional connectivity and cognitive function in mild to moderate Alzheimer’s disease. Fifty-eight participants were randomised to receive a course of either active or sham iTBS. Stimulation was applied to four brain sites sequentially in each treatment session: left DLPFC, right DLPFC, left PPC, and right PPC. Neurobiological (EEG), cognitive (CogState, ADASCog), and functional (QoL-AD, GDS) assessments were undertaken at baseline and end of treatment. Cognitive and functional assessments were also conducted at 3-months (blinded) and 6-months (active group only) following end of treatment. Active iTBS resulted in a significant and large increase in resting state gamma connectivity as well as improved delayed recall on an episodic memory task. Both baseline gamma connectivity, and change in gamma connectivity, were found to be predictive of improved delayed recall following active treatment. These findings support future research into iTBS for the treatment of Alzheimer’s disease focussing on protocol optimisation.

## 1. Introduction

Alzheimer’s disease is a neurodegenerative cognitive disorder and the most common form of dementia. According to the World Health Organisation (WHO) there are currently an estimated 55 million people with dementia worldwide. Without a significant treatment breakthrough, this number is predicted to increase to 78 million by 2030 and 139 million by 2050 (WHO, 2021). There are currently six FDA approved pharmacological treatments for Alzheimer’s (Alzheimer’s Association, 2021). Five of the six target the cholinergic and/or glutamatergic systems and have been shown to produce limited symptomatic benefits, not impact disease course and possess side effects limiting tolerability (Alzheimer’s Association, 2021; Husna Ibrahim et al., 2020). In June 2020 the FDA provided accelerated approval to aducanumab, an amyloid beta-directed monoclonal antibody and the first disease modifying therapy (Lalli et al., 2021). Approval was given despite the fact that although aducanumab was shown to reduce amyloid burden it was not associated with clinical improvement (Lalli et al., 2021; Mullard, 2021; Schneider et al., 2019). Treatment approaches which can modulate proximate therapeutic targets, and hence have a greater likelihood of producing clinical improvement, are still urgently needed (Canter et al, 2016).

There is growing evidence that neural network dysfunction is a likely proximate cause of cognitive impairment in Alzheimer’s disease, with accumulation of disease protein associated with disrupted connectivity in function-critical neural networks such as the frontoparietal network (FPN) and the default mode network (DMN) (Canter et al, 2016; Contreras et al., 2019; Damoiseaux et al., 2012; Grothe et al., 2016; Hasson et al., 2017; Paplop et al., 2010; Paplop et al., 2016). Changes in functional connectivity with Alzheimer’s have been reported across imaging modalities, including functional Magnetic Resonance Imaging (fMRI), electroencephalography (EEG) and magnetoencephalography (MEG) (Babiloni et al., 2018; Bagattini et al., 2019; Briels et al., 2020; Contreras et al., 2019; Damoiseaux et al. 2012; Engles et al., 2015; Madal et al., 2018). With the direction of these connectivity changes (i.e. increases or decreases) appearing to be dependent upon frequency band, brain region/network, and stage of illness (Babiloni et al., 2018; Briels et al., 2020; Engles et al., 2015). Critically, however, *dys*functional connectivity (i.e. increases or decreases in connectivity) in Alzheimer’s has been shown to be associated with cognitive symptoms (Contreras et al., 2019; Damoiseaux et al. 2012; Engles et al., 2015; Hoy et al., 2022).

It has been suggested that this dysconnectivity may be a result of impairments in inhibitory interneuron function and subsequent excitatory-inhibitory imbalance, particularly between anterior and posterior regions (Bagattini et al., 2019; de Haan et al., 2017; Paplop et al., 2010; Paplop et al., 2016; Pievani et al., 2014). In a recent study we provided some experimental support for this, finding that higher frontoparietal theta connectivity in people with Alzheimer’s disease was significantly associated with reduced prefrontal cortical activation (assessed using EEG combined with Transcranial Magnetic Stimulation [TMS-EEG]). We also found that higher theta connectivity was associated with poorer episodic memory (Hoy et al, 2022). This analysis was conducted from the baseline data of a subset of participants from the clinical trial which is the focus of the current paper. Collectively, these findings provide support for the investigation of a treatment approach which targets impaired cortical activity in order to restore optimal connectivity and improve cognitive function. Non-invasive brain stimulation techniques are particularly well suited to such an approach.

To date there have been less than a dozen previous sham-controlled trials investigating Transcranial Magnetic Stimulation (TMS) for Alzheimer’s disease, the majority of which have demonstrated small to moderate effects on cognition using standard TMS approaches (Chu et al., 2021; Liu et al., 2021). Despite these promising results, there remain limitations with past research. In particular, none of these trials have used tailored brain stimulation approaches to optimally target large scale network dysfunction (Chu et al., 2021; Liu et al., 2021). Direct targeting of proposed therapeutic targets, and measurement of successful target engagement, is critical for the efficient and effective development of novel treatment approaches. In addition, the majority of the trials conducted to date have been in small samples (the vast majority were n<30, with only 2 trials having n>50) and none have provided full treatment doses to multiple brain regions as is likely to be required in Alzheimer’s disease (all have applied stimulation to only one brain site per session or multiple sites but on alternating treatment days) (Chu et al., 2021; Liu et al., 2021). Intermittent Theta Burst Stimulation (iTBS) is a ‘patterned’ form of stimulation that is thought to more closely mimic endogenous brain activity and has been shown to modulate frontoparietal functional connectivity (Hoy et al., 2016). iTBS offers a number of advantages over ‘standard TMS’ which are of particular relevance to a therapeutic approach for Alzheimer’s (Chung et al., 2016). The most striking difference between iTBS and standard TMS is administration time. A typical TMS treatment protocol takes approximately 40 minutes whereas iTBS is complete within 3 minutes, meaning that iTBS allows for stimulation of multiple brain regions in every treatment session (Chung et al., 2016). To date, there have been no clinical trials of iTBS for the treatment of Alzheimer’s disease.

In the current study we sought to investigate iTBS applied to multiple brain locations in the treatment of patients with Alzheimer’s disease in a randomised controlled clinical trial. We provided iTBS to highly connected heteromodal hubs of networks known to be dysfunctional in Alzheimer’s disease (i.e. the prefrontal cortex and posterior parietal cortex) in an attempt to directly restore neuronal function and thus optimal connectivity. Our primary hypotheses were that active iTBS, compared to sham, would significantly modulate resting state functional connectivity, and improve performance on an episodic memory task following a 6-week treatment course. We also conducted exploratory analyses to investigate (1) the relationship between resting state functional connectivity and episodic memory outcomes and, (2) the impact of the treatment course on broader cognitive and functional outcomes.

## 2. Materials and Methods

### 2.1 Trial Protocol

This was a double-blind parallel randomized sham-controlled trial. Participants were randomised to receive 21 sessions of either active or sham iTBS over a six-week treatment course. Both raters and participants were blind to the treatment group. While the research nurses administering treatments were aware of the participants’ treatment groups, they were not involved in any cognitive, functional or EEG assessments and were counselled to avoid discussion of treatment related aspects with participants which may compromise blinding. Randomisation occurred via the generation of a random number sequence and participants were randomised prior to their first treatment session. Cognitive, functional and EEG assessments were conducted at baseline and at the end of treatment. A brief cognitive assessment was also conducted mid treatment course (i.e. week 3). Follow up cognitive and functional assessments were conducted at 3-months (blinded) and 6-months (active group only) after the end of treatment. See Supplementary Figure One for full study design. Ethics approval was granted by Monash University and the Alfred Health ethics committees. Written consent was obtained prior to undertaking any study procedures.

### 2.2 Participants

A total of 58 participants with mild to moderate Alzheimer’s disease were recruited to the study, 2 participants withdrew prior to completing all baseline assessments. See Figure One for the CONSORT flow diagram and Table One for demographic and clinical characteristics.

**Figure One:**
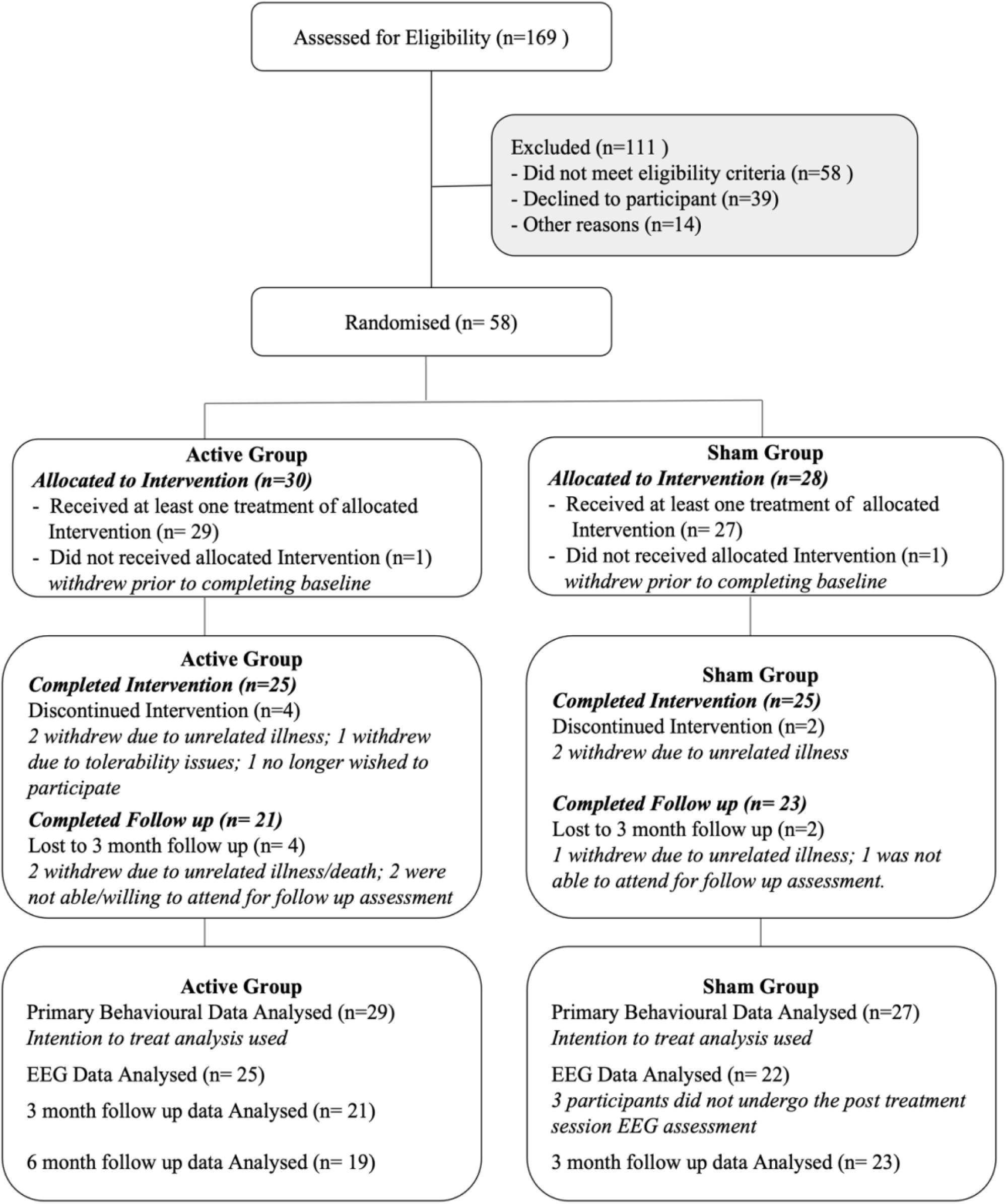
CONSORT Diagram.

**Table One.**
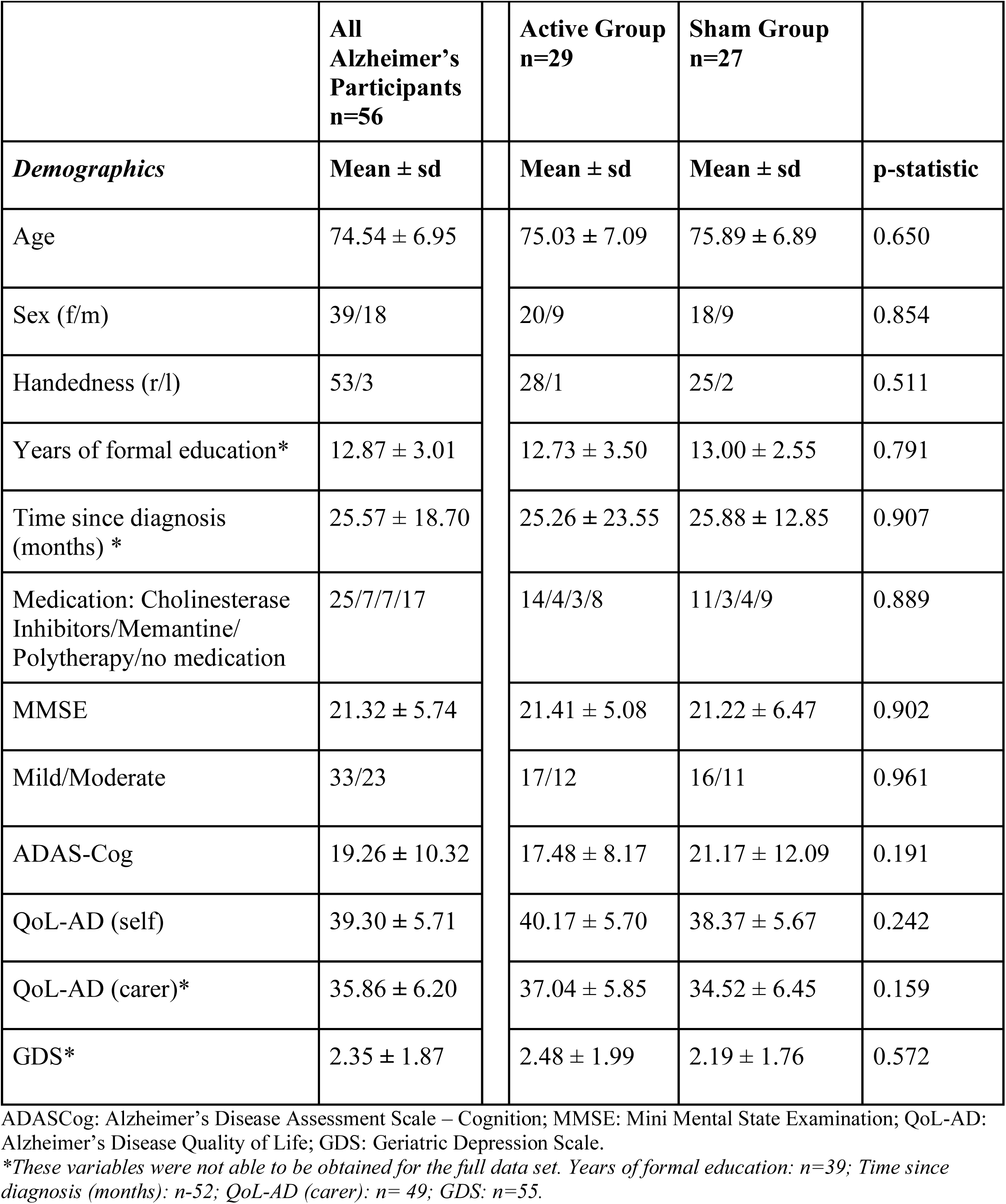
Demographics and clinical characteristics for Alzheimer’s participants (combined and by treatment group).

Inclusion criteria included a diagnosis of dementia of the Alzheimer’s type based on general medical, neurological, and neuropsychological examinations according to the National Institute of Neurological and Communication Disorders–Alzheimer’s Disease and Related Disorders Association (NINCDS–ADRDA) criteria for probable Alzheimer’s disease and the DSM-IVTR clinical criteria for dementia of the Alzheimer’s type (First et al., 2002; McKhann et al. 2011). Diagnosis was provided by the referring clinician. Participants were required to have a score of at least 10 on the Mini-Mental State Evaluation (MMSE) (Folstein et al., 1978). Participants were excluded if they had a history of any concomitant major and unstable neurological or serious medical conditions (including psychiatric); metal in the cranium, cochlear implant, medication pump or other electronic device; a DSM-IV history of substance abuse or dependence in the last 6 months; or had a history of seizures.

Participants were required to either not be taking psychotropic medication or their dose of medication be unchanged for a minimum of 4 weeks prior to entry into the study and throughout their involvement. A total of 39 out of 56 participants were taking psychotropic medication. Of the 32 participants on monotherapy 25 were taking cholinesterase inhibitors (20 donepezil, 4 galantamine, 1 rivastigmine) and 7 were on memantine. There were 7 participants on polytherapy (1 donepezil + memantine; 1 rivastigmine + memantine; 1 donepezil + citalopram; 1 donepezil + clonazepam; 1 donepezil + sertraline; 1 memantine + duloxetine; 1 galantamine + memantine). Seventeen participants were not taking any psychotropic medication.

Please note, we have previously conducted an analysis of the baseline data from this trial in a subset of participants (n=40), to investigate potential neurophysiological markers of symptom severity (Hoy et al., 2022).

### 2.3 Intervention: intermittent Theta Burst Stimulation

iTBS treatment was provided using a Neurosoft MS/D (Neurosoft, Ivanovo, Russia) magnetic stimulator with a cooled angulated figure-of-8 coil (Coil winding diameter – 2×100 mm). Stimulation was applied to four brain sites sequentially at each treatment (i.e. left DLPFC, right DLPFC, left PPC, right PPC always in the same order). All sites were stimulated using 3-pulse 50-Hz bursts applied at 5 Hz with a 2-second train of stimulation repeated every 10 seconds for a total of 180 seconds per site (i.e. 600 pulses). iTBS was applied at 100% of the Resting Motor Threshold which was determined bilaterally using standard published methods (Fitzgerald et al., 2002). Treatment sites were determined using the international 10-20 system of measurement (Herwig et al., 2003), namely Left DLPFC: F3; Right DLPFC: F4; Left PPC: P3; Right PPC: P4. Each participant had these sites measured-up and marked on a close-fitting cloth cap prior to their first treatment. This personalised cap was then used for all treatments to ensure consistent coil placement. The treatment coil was positioned at the individually defined treatment site with the handle pointing back and away from the midline at a 45-degree angle and tangential to the scalp. Sham stimulation was applied using identical treatment parameters but with the coil angled at 90 degrees off the head.

### 2.4 Assessments

See Supplementary Figure One for full study design.

#### 2.4.1 Resting state-EEG

EEG data was obtained from a 40-scalp electrode montage in the standard 10-20 positions (Quickcap, Compumedics Ltd., Australia). The cap was positioned such that FPz was located at 10% of the distance from the nasion to inion above the nasion. Electrode impedances were regularly checked between recordings and kept below 5 kΩ throughout the experiment. The EEG signal was amplified (10000x), filtered (DC-3500 Hz) and digitized (10 kHz; Synamps2, Compumedics Ltd.) and recorded on a computer for offline analysis. EEG was recorded while participants were at rest, in both an eyes open (3 mins) and eyes closed (3 mins) condition. See Supplementary Material for details of electrodes used, pre-processing and analysis of rs-EEG data. Resting state EEG was recorded at the baseline and end of treatment assessments.

#### 2.4.2 Cognitive and Functional

The primary cognitive outcome measure, episodic memory, was measured using the International Shopping List (ISL) task from the CogState Alzheimer’s battery (Maruff et al., 2013). Episodic memory was chosen as it is strongly associated with frontal-parietal brain activity (Contreras et al., 2019) and impairment in Alzheimer’s disease is evident even in the early stages of the illness (Snyder et al., 2014). We specifically looked at delayed verbal episodic memory (ISL Delayed Recall) and immediate verbal episodic memory (ISL Total).

We additionally assessed cognition using the ADASCog (Connor and Sabbagh., 2008). The ADASCog is a specialised Alzheimer’s cognitive assessment made up of 11 tasks across the cognitive domains most often impacted by Alzheimer’s, with scores from 0-70 where higher scores indicate poorer cognitive performance. To assess quality of life we used the Quality of Life in Alzheimer’s Disease (QoL-AD) self and carer versions (Logsdon et al., 2002). Finally, we also used the Geriatric Depression Scale (GDS) to assess depression symptomatology (Yesavage, 1988). All cognitive and functional measures were conducted at baseline, end of treatment, at the 3-month follow up and at the 6-month follow-up (active group only). The International Shopping List (ISL) task was additionally conducted during the treatment course at the week 3 review.

### 2.5 Statistical Analysis

EEG data were pre-processed using the extreme outlier rejection and wavelet enhanced independent component analysis artifact reduction methods from RELAX, a cleaning pipeline developed using EEGLAB and Fieldtrip functions within MATLAB (Bailey et al. in preparation; Brunner et al., 2013; Delmore and Makeig, 2004; Oostenveld et al., 2011) (2019). Statistical analyses were performed using the Network Based Statistic (NBS) (Zalesky et al., 2010) (2010), and SPSS version 24 (IBM Corp, Armonk, NY, USA. The analysis we undertook for our primary and exploratory hypotheses are described in detail below. A significance value of p<0.05 was used unless otherwise specified.

#### 2.5.1 Resting State EEG analysis: Functional Connectivity

Between group differences (active, sham) in resting state functional connectivity at the 6-week timepoint (i.e. following the iTBS treatment course) were analysed using NBS via non-parametric cluster-based permutation methods, with mass univariate independent sampled t-tests which were controlled for multiple comparisons using cluster statistics. The first step in NBS is to run univariate statistical tests (in the case of the present study, *t*-tests) at each node-node connection. Test statistics exceeding an a priori determined critical *t*-value threshold are then admitted to a set of suprathreshold connections. Suprathreshold node-node connections that are topologically connected to each other form connected networks. These networks are then submitted to between-group statistical analysis at the network level by attributing family-wise error corrected p-values to each group cluster. These family-wise error corrected p-values were derived using 5000 randomised permutations of the cluster level *t*-statistics to derive the null-distribution of no effect, and thus the adjusted p-value. Please refer to (Zalesky et al., 2010) for more information regarding the application and methodologies of NBS. Separate analyses were run on the wPLI values for each frequency band of interest (theta and gamma) in NBS. An *a priori* critical *t*-value threshold was set at 2.3948 (df = 46) for individual pairs of electrodes, corresponding to a p-value of 0.01. The FWER corrected alpha threshold for testing of connected clusters was specified at alpha = 0.05. For networks identified as differing between the active and sham condition at the endpoint recording using the NBS, strength of connectivity within the significant network was extracted and averaged for all participants at the baseline and end of treatment timepoints for subsequent analysis.

#### 2.5.2 Cognitive and Functional Outcomes

T-tests and χ 2 tests were used to examine differences between the groups on demographic and baseline variables (See Table One). Intention to treat analyses were conducted for all cognitive and functional variables using linear mixed models (LMM). Data were checked for normality of residuals, homoscedasticity and presence of outliers; data was also assessed for the missing at random (MAR) assumption which was met. The data showed only minor violations of normality and due to the robustness of mixed-effects models to minor violations we proceeded with the LMM analysis (Schielzeth et al, 2020). The unstructured covariance structure and maximum likelihood (ML) were used to estimate parameters with Satterthwaite approximation to degrees of freedom (Barton and Peat, 2014).

For the primary cognitive outcome measure LMMs were conducted with ISL Delayed Recall and ISL Total as dependent variables, with fixed effects of group (active, sham) and time (baseline, week 3, end of treatment). We also conducted LMMs for our exploratory analyses looking at effects on broader measures of cognition and function, namely for ADASCog, QoL-AD (self, carer) and GDS, with fixed effects of group (active, sham) and time (baseline, end of treatment). The 3 month and 6 month follow up data were also analysed using LMMs. These results are reported in full in the Supplementary materials.

#### 2.5.3 Regression Analysis

Multiple regression analyses were conducted to investigate the relationship between resting state functional connectivity and episodic memory outcomes. Any change scores used were calculated as post-pre. Regressions were run separately for the active and sham groups.

## 3. Results

### 3.1 Resting State-EEG: Functional Connectivity

Network based statistics identified a network exhibiting greater gamma connectivity in the active group compared to the sham group at end of treatment in the eyes closed condition (p = 0.055), See Figure Two. The identified network encompassed frontal, parietal, and occipital regions both within and between hemispheres; with predominantly frontal to frontal connectivity and frontal to parietal/occipital connectivity. There were no other changes in network activity.

**Figure Two.**
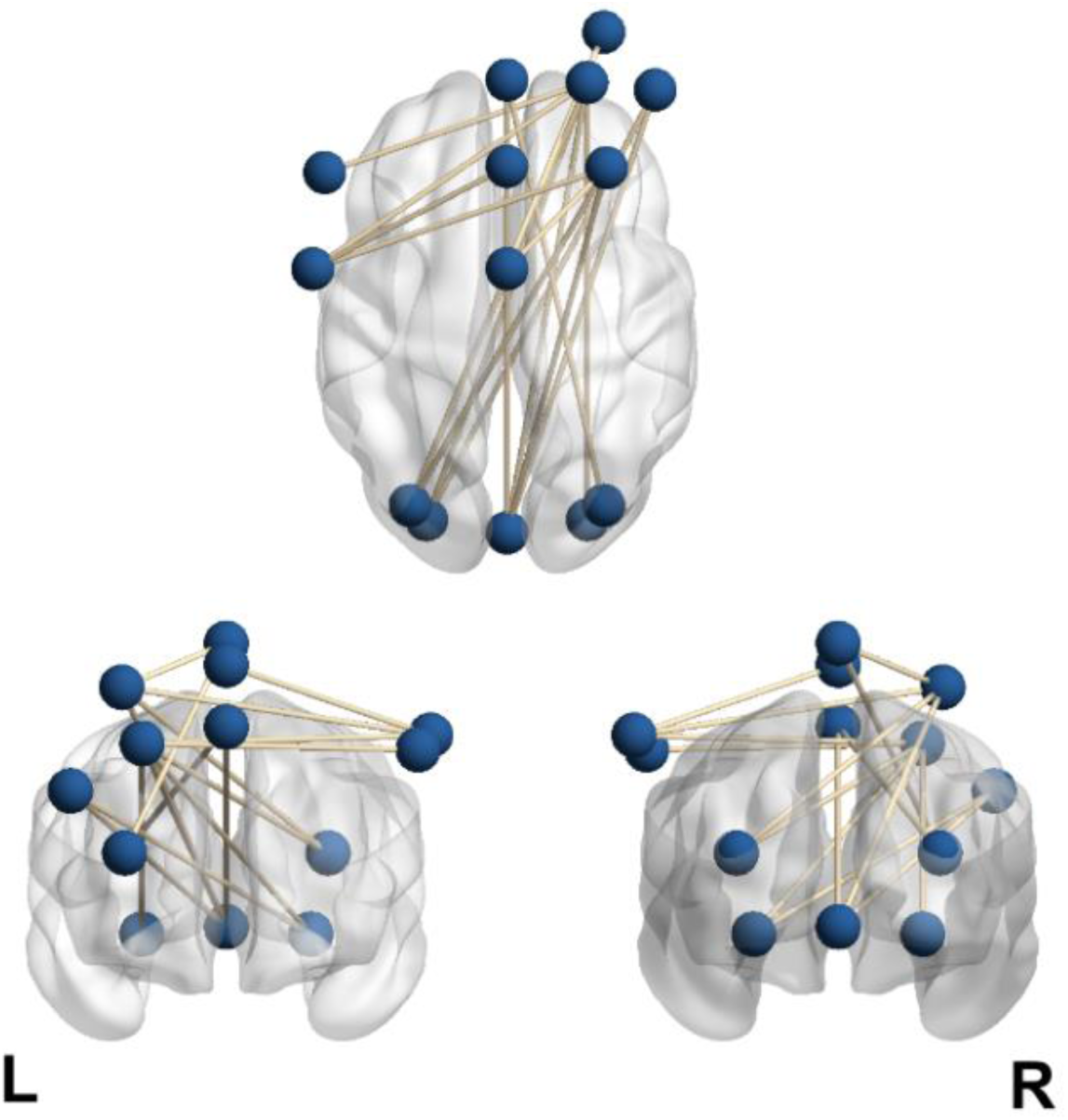
Weighted Phase Lag Index (wPLI) network connectivity map between significant electrodes reflecting greater gamma connectivity in the active group compared to the sham group at end of treatment. Grey bars reflect significant connectivity between pairs of electrodes. wPLI values were derived from EEG data obtained while participants were seated at rest, eyes closed. NBS identified a gamma network using a primary multiple comparison control threshold for significance between pairs of electrodes of 0.01. The network was visualised using the BrainNet Viewer (Xia et al., 2013).

In light of the stringent primary alpha threshold for connectivity comparisons using the NBS (p=0.01) and near significant differentiation between active and sham groups, averaged values from the gamma network were extracted for further analysis. The data showed minor normality violations and only a single extreme outlier (i.e. z value of greater than 3.29 in a participant in the active group at baseline). Therefore, due to its robustness, we conducted an ANOVA test of significance. We ran the analysis with and without the outlier and as there was no difference in the significance found we report here the analysis with the outlier retained. Means and standard deviations for averaged gamma wPLI values are provided in Supplementary Table One.

There was a significant effect of Time (F_(1, 46)_= 4.329, *p*= 0.043), whereby there was a significant increase in strength of averaged gamma connectivity from baseline to end of treatment. There was also a significant Time by Treatment Group interaction (F_(1, 46)_ = 14.256, *p* <0.001). Post hoc analyses were conducted to explore the interaction. There were no differences in averaged gamma connectivity between the active and the sham groups at baseline (F_(1, 46)_= 0.00, *p* = 0.984). There was, however, a significant, and large, difference between the groups at the end of treatment (F_(1, 46)_= 12.805, *p* =0.001, *d*=1.05), with the active group showing significantly greater averaged gamma connectivity than sham. There was significant, and large, increase in averaged gamma connectivity from baseline to end of treatment in the active group (t_(24)_ = 3.618, *p* =0.001, *d*=0.724), while there was no change in the sham group (t_(22)_ = 1.517, *p* =0.144). See Figure Three.

**Figure Three.**
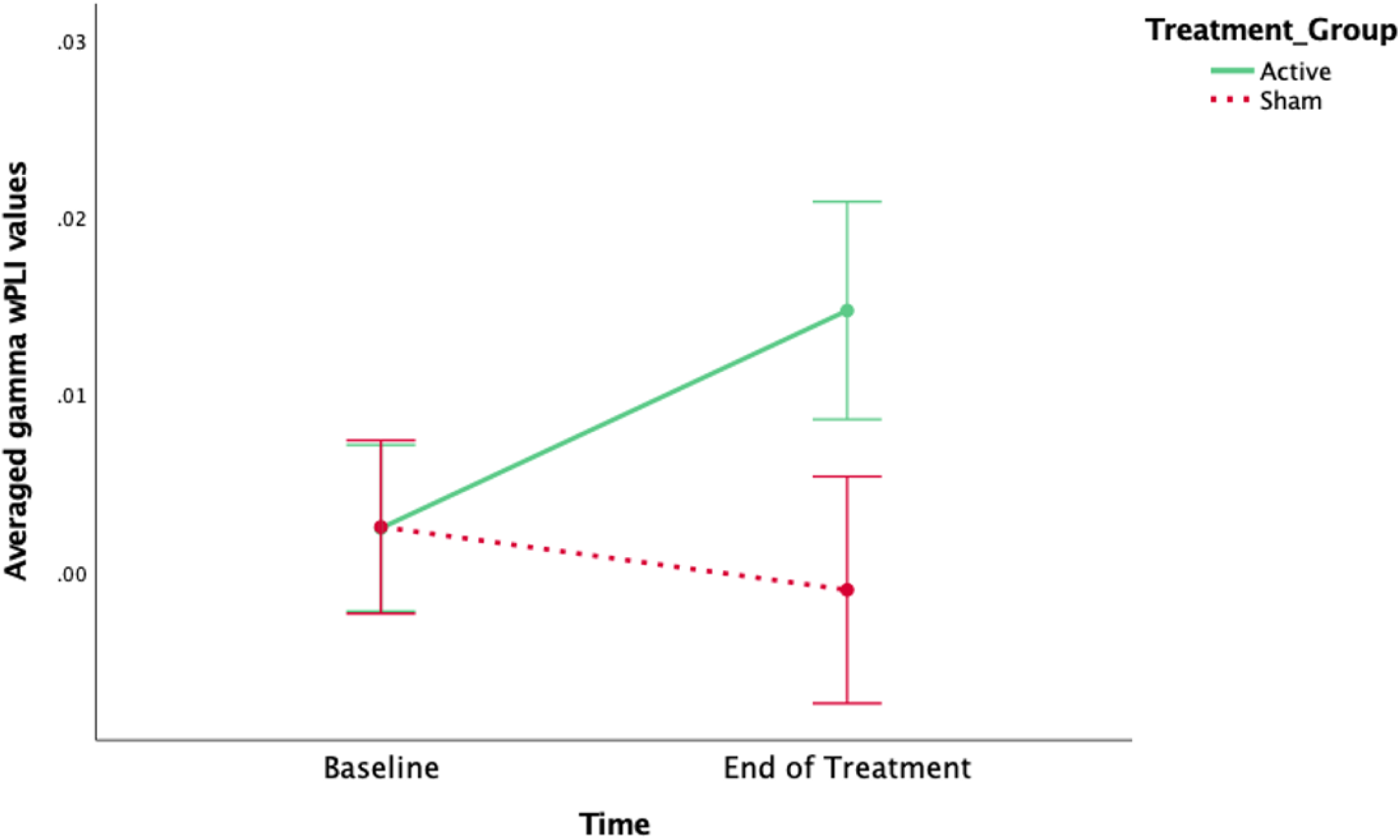
Averaged gamma wPLI values from the NBS identified network over time as a function of the treatment group.

### 3.2 Cognitive and Functional Outcomes

Means and standard deviations for all cognitive and functional outcomes across all timepoints as a function of treatment group are provided in Supplementary Table Two.

#### 3.2.1 Primary Cognitive Outcomes

The LMM for the ISL Delayed Recall showed no main effect of Time (F _(2, 51.100)_ = 2.625, p = 0.082), there was however a significant Time by Treatment Group interaction (F _(2, 51.024)_ = 3.497, p = 0.038). Post-hoc analyses revealed a significant improvement in the active group from Baseline to End of Treatment (mean difference = 0.611, p = 0.036) and a significant decline in the sham group from Week 3 to End of Treatment (mean difference = -0.814, p = 0.018). There were no significant differences between the groups at any time point (baseline: mean difference = 0.103, p = 0.814; week 3: mean difference = -0.144, p = 0.774; end of treatment: mean difference = 0.932, p = 0.073). See Figure Four.

**Figure Four.**
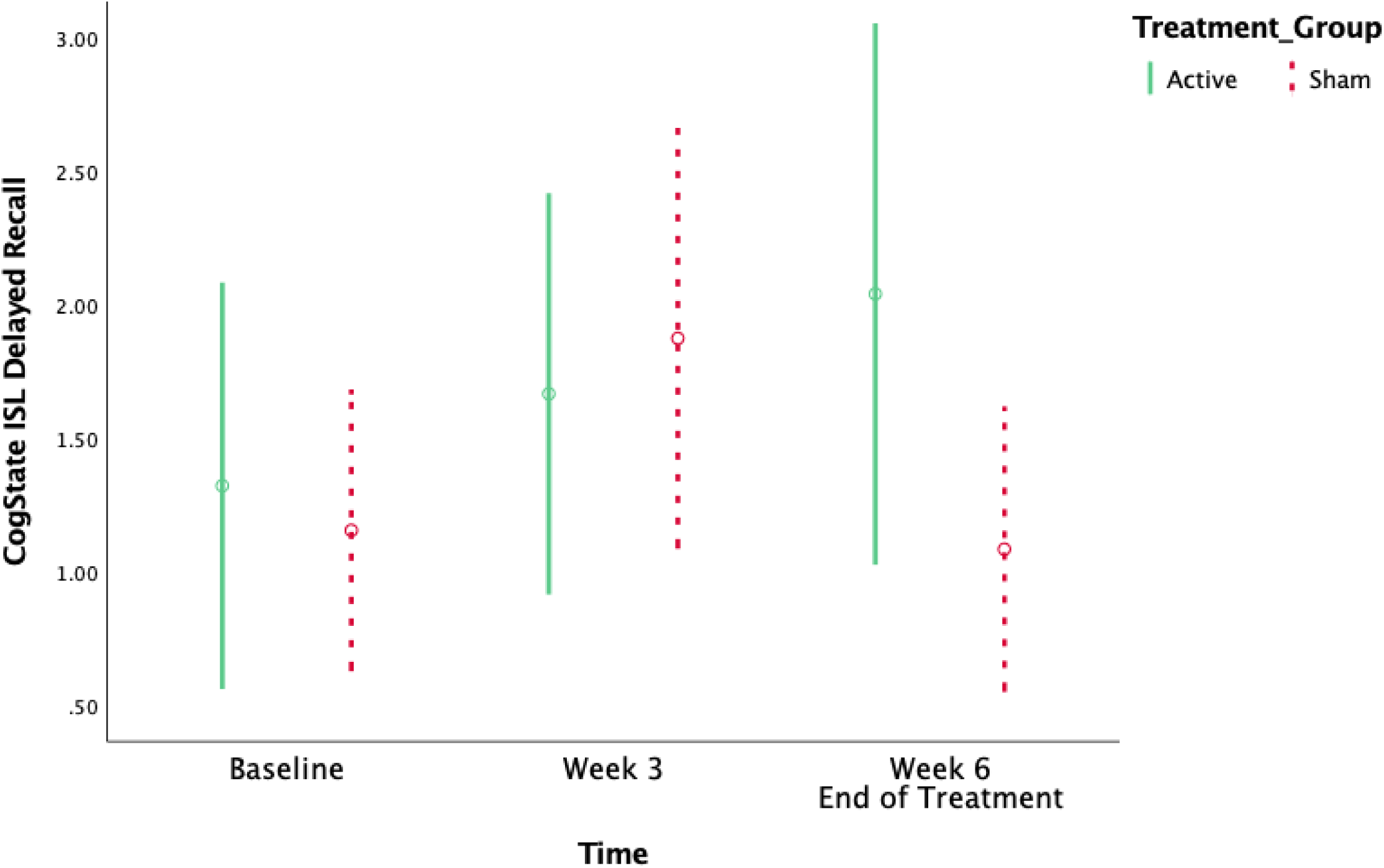
CogState ISL Delayed Recall mean scores over time as a function of treatment group.

For the ISL Total scores there was a near significant main effect of Time (F _(2, 51.024)_ =3.130, p = 0.052) and no interaction effect (F _(2, 51.024)_ =0.175, p = 0.840). Post-hoc analyses to further clarify the main effect of Time showed a significant improvement, irrespective of treatment group, from Baseline to End of Treatment (mean difference = 1.307, p = 0.018).

#### 3.2.2 Exploratory Outcomes

There were no significant main or interaction effects for the ADASCog, QoL-AD (self, carer) and GDS, See Table Two.

**Table Two.** Linear Mixed Models of Exploratory Clinical and Functional Outcomes (Baseline to End of Treatment).

| Assessment | Effect over Time | p value | Time by Group Effect | p value |
| --- | --- | --- | --- | --- |
| ADASCog | $F(1, 50.005) = 0.523$ | 0.473 | $F(1, 50.005) = 0.507$ | 0.480 |
| QoL-AD self | $F(1, 49.994) = 2.222$ | 0.142 | $F(1, 49.994) = 0.167$ | 0.685 |
| QoL-AD carer | $F(1, 32.789) = 0.238$ | 0.629 | $F(1, 32.789) = 0.382$ | 0.541 |
| GDS | $F(1, 49.754) = 0.033$ | 0.858 | $F(1, 49.754) = 2.504$ | 0.120 |

ADASCog: Alzheimer's Disease Assessment Scale – Cognition; QoL-AD: Alzheimer's Disease Quality of Life; GDS: Geriatric Depression Scale.

### 3.3 Regression

The relevant assumptions (i.e. linearity of relationship between IVs and DV, multicollinearity; independence, homeoscedasctity and normality of residuals; no extreme outliers) were checked and met. Hierarchical linear regression analysis was conducted to evaluate the prediction of *change in delayed recall* from *baseline gamma connectivity, change in gamma connectivity*, and *baseline delayed recall* (included as a control). In the first block analysis, the predictor variables *baseline gamma connectivity*, and *baseline delayed recall* (included as a control) were analysed, for the second block the *change in gamma connectivity* predictor was added to the analysis.

For the active group the results of the first block analysis revealed a significant model (F_(2,22)_ = 6.037, *p*=0.009, R^2^ = 0.376, R^2^_Adjusted_ = 0.314) which explained 37.6% of the variance in change in delayed recall. From the model the only significant predictor was baseline gamma connectivity (Beta =0.581, t_(22)_ = 3.267, *p*=0.004), with a unique contribution of r= 0.333.

In the second block, where change in gamma connectivity was added as a predictor variable, the model was again significant (F_(3,22)_ = 7.376, *p*=0.002, R^2^ = 0.538, R^2^_Adjusted_ = 0.465) and accounted for a significant amount of additional variance in change in delayed recall (F Change_(1,19)_= 6.647, *p*=0.018, R^2^ Change= 0.162). Significant predictors from the model were baseline gamma connectivity (Beta =0.715, t_(22)_ = 4.321, *p*<0.00; unique contribution of r = 0.454) and change in gamma connectivity (Beta = 0.425, t_(22)_ = 2.578, *p*=0.018; unique contribution of r = 0.162). See Table Three.

**Table 3.**
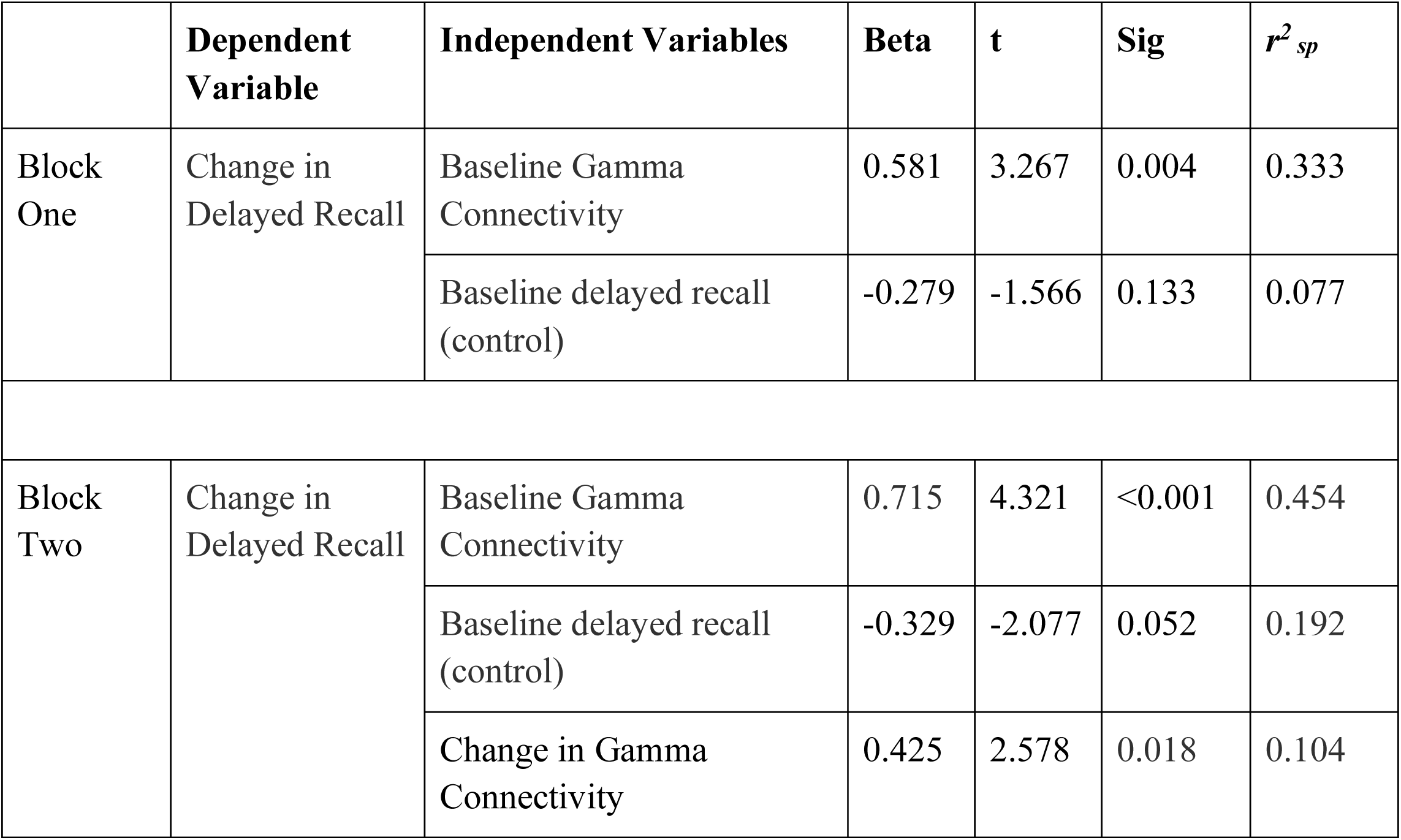
Hierarchical Linear Regression Analysis Coefficients for Active Group

For the sham group neither block one (F_(2,20)_ = 2.671, *p*=0.096, R^2^ = 0.229, R^2^_Adjusted_ = 0.143) or block two (F_(3,20)_ = 1.686, *p*=0.208, R^2^ = 0.229, R^2^_Adjusted_ = 0.093) revealed a significant model. See Table Four.

**Table 4.** Hierarchical Linear Regression Analysis Coefficients for Sham Group

| | Dependent Variable | Independent Variables | Beta | t | Sig | $r^2_{sp}$ |
| --- | --- | --- | --- | --- | --- | --- |
| Block One | Change in Delayed Recall | Baseline Gamma Connectivity | 0.099 | 0.444 | 0.662 | 0.008 |
|  |  | Baseline delayed recall (control) | -0.507 | -2.270 | 0.036 | 0.221 |
| Block Two | Change in Delayed Recall | Baseline Gamma Connectivity | 0.114 | 0.408 | 0.689 | 0.008 |
|  |  | Baseline delayed recall (control) | -0.516 | -2.055 | 0.056 | 0.192 |
|  |  | Change in Gamma Connectivity | 0.025 | 0.95 | 0.925 | <0.001 |

Overall, this indicates that in the active treatment group the predictors of change in delayed recall were baseline gamma connectivity (i.e. higher gamma connectivity at baseline predicting greater change in delayed recall) and change in gamma connectivity (i.e. greater change in gamma connectivity following active treatment predicting greater change in delayed recall).

### 3.4 Tolerability and Blinding

There was no significant difference in the reporting of side effects between the two groups (*x*^*2*^ (1, 55) =1.002, p = 0.317). From the active group, 3 participants reported experiencing side effects (i.e. headache, site discomfort), compared to 1 participant in the sham group. Of the 4 participants who reported side effects 3 reported issues with tolerability of the treatment (2 in active group, 1 in sham group). Again, there was no significant difference in tolerability between the two groups (*x*^*2*^ (1, 55) 0.315, p = 0.574). There were no treatment related serious adverse events in either group. From enrolment to the end of the 6-week treatment course 8 participants withdrew from the study (13.8% attrition during intervention phase), from end of treatment course to the 3 month follow up 6 participants withdrew from the study (12% attrition during the blinded follow up phase); see Figure One for more detail.

Due to a data collection error we are only able to report on blinding fidelity from a subset of the sample, approximately half of the total sample. Blinding assessment was conducted at the end of the treatment course for participants, raters and carers. The number of correct guesses as to which treatment group participants were in did not significantly differ from chance for any of the groups (participant: *x*^*2*^ (1, 28) 0.000, p = 1.00; rater: *x*^*2*^ (1, 29) 0.056, p = 0.812; carer: *x*^*2*^ (1, 22) 0.105, p = 0.949).

## 4. Discussion

The current study sought to investigate whether iTBS could successfully engage a proposed proximate therapeutic target in Alzheimer’s, namely dysfunctional neural connectivity. The primary hypotheses, that iTBS would significantly modulate resting state connectivity and improve episodic memory, were supported. Specifically, a 6-week treatment course of iTBS significantly enhanced gamma connectivity and improved delayed recall for episodic memory. The active iTBS group showed a significant and large enhancement of gamma connectivity across a widespread network encompassing frontal, parietal, and occipital regions both within and between hemispheres. There was no change in gamma connectivity seen across this network in the sham group. Similarly, there was a significant improvement in delayed recall in the active group from baseline to end of treatment, while in sham there was a significant decline from week 3 of treatment to end of treatment following an initial non-significant improvement. Critically, in the active treatment group both strength of gamma connectivity at baseline and increased gamma connectivity following treatment were significant predictors of change in delayed recall. An increase in total episodic memory (i.e., total number of words learned across three trials) from baseline to treatment end irrespective of group was also found, indicating that the improvement in delayed recall in the active group could not be attributed to differences in total learning between the groups. There was no effect of treatment on the broader cognitive and functional measures, namely the ADASCog, QoL (self, carer) or the GDS.

The current trial provides the first evidence of successful engagement of a therapeutic target, i.e. dysfunctional neural connectivity, with iTBS in the treatment of Alzheimer’s. While there was an improvement in delayed recall, there were no significant improvements seen on broader measures of cognition, namely the ADASCog. A number of previous trials of TMS for Alzheimer’s have reported improvements on the ADASCog, however all have either included cognitive training and/or a longer daily treatment duration than the current trial (i.e. 5 treatments a week for 5 or 6 weeks) (Lee et al., 2016; Sabbagh et al., 2021; Wu et al., 2015; Zhao et al., 2017). In the current trial daily (Mon-Fri) treatments were provided for 3 weeks, followed by a 3-week maintenance schedule of a progressively reducing number of treatments per week (3, 2, 1). The maintenance schedule aimed to provide ‘consolidation’ of any improvements seen during the daily treatment phase and was largely informed by our previous TMS clinical trial work in depression (Fitzgerald et al., 2013). While treatment was applied to four brain regions in every treatment session, thus providing a higher treatment dose per session than past trials, it is possible that the duration of daily treatment was not long enough to produce broader cognitive improvement in the current trial. Indeed, it is likely that for a progressive illness such as Alzheimer’s it is the duration and frequency of treatments that is the most critical factor with respect to dosing. Similarly, the lack of significant functional improvement is likely to be due to the relatively short duration of treatment.

The findings of enhanced gamma connectivity following iTBS are consistent with a recent study of TMS in Alzheimer’s showing that stimulation enhanced gamma power and promoted functional integration and effective connectivity (measure via fMRI and TMS-EEG respectively) (Liu et al., 2021). Gamma connectivity in particular has been implicated in critical cognitive processes (such as learning, attention and memory) and is an important mechanism for integrating activity within and between neural networks during cognitive processing (Bosman et al., 2014; Kaiser and Lutzenberger et al., 2005). The deterioration of cognition due to the dysregulation of functional connectivity in Alzheimer’s disease, including gamma connectivity, is supported by past research (deHann et al., 2009; Contreras et al., 2019; Damoiseaux et al. 2012; Engles et al., 2015). Importantly, the results of the regression analyses in the current study further supports the role of gamma connectivity as a key therapeutic target, showing that in the active treatment group stronger gamma connectivity at baseline and increased gamma connectivity following treatment were significant predictors of improved delayed recall. This would indicate that a degree of ‘intact’ gamma connectivity at baseline is needed for iTBS to subsequently enhance connectivity, and that the degree of enhancement achieved with iTBS predicts the degree of cognitive improvement.

The ability of iTBS to modulate a pathophysiological marker of Alzheimer’s is significant and provides strong impetus for future research into protocol optimisation. In particular, as discussed above, in light of the progressive nature of Alzheimer’s disease, longer treatment durations with iTBS should be explored. In addition, there is a strong case for investigating the combination of iTBS with disease modifying therapies such as aducanumab. While reduction of amyloid burden may be important for halting disease progression, it will not restore the damage already done to the neuronal networks that are crucial for successful cognition. Conversely, while iTBS is unlikely to be able to alter the course of the disease, the current results suggest that it can restore neuronal function to an extent. The combination of these approaches may be the best chance we have of developing a comprehensive approach which can halt disease progression and restore function.

The findings of the current study should be interpreted in the context of the following limitations. While the sample size was larger than the majority of past TMS trials in this patient group, it can still be considered statistically small. This limited the ability to conduct subgroup analyses, which might have included an exploration of the impact of illness severity (mild, moderate), or an analysis including response and non-response, as has been done previously (Sabbagh et al., 2020). Future research should also look to more comprehensively characterise participants using blood biomarkers and genotyping, whose validity and availability have increased in the time since this trial was commenced. Similarly, we suggest more comprehensive assessment of functional connectivity outcomes including the additional use of fMRI and task related assessments for future work in the area. Finally, we used the international 10-20 system of measurement to localise iTBS treatment sites rather than neuronavigation. While the 10-20 system of measurement has been shown to be an acceptable method of TMS treatment site localisation (Fitzgerald et al., 2009), MRI based localisation methods should be utilised where possible and funding allows.

The conceptualisation of Alzheimer’s as a disorder of connectivity coupled with the very limited success rate of treatments developed to date strongly supports the investigation of alternative approaches such as brain stimulation. The current study showed that iTBS was able to successfully engage the proposed therapeutic target and that this was predictive of improved cognition. This is a critical initial step in treatment development. These findings provide strong support for the future investigation of iTBS for the treatment of Alzheimer’s in larger samples and with longer treatment durations; as well as the consideration of combining iTBS with disease modify therapies to maximise the efficacy of both approaches.

## Data Availability

At the time of ethical approval of this study data access was not specifically addressed (this is now done as a matter of course). As such research participants of this
study did not agree for their individual data to be shared publicly. The corresponding author will apply to the relevant ethics bodies to request permission for the
open access sharing of de-identified data via a public repository (i.e. Monash University Figshare).

## Acknowledgements

KEH was supported by National Health and Medical Research Council (NHMRC) Fellowships (1082894 and 1135558). PBF was supported by an Investigator Fellowship from the NHMRC (1193596). This research was supported by project funding from the NHMRC (1082894), the Mason Foundation (MAS2015F011 & MAS2017F003), State Trustees Australia Foundation (STAF-MR-2018-01), a Monash University Strategic Grant (SGS16-0305) and the community of Monash University alumni donors. We also acknowledge the time, involvement and feedback from the participants and their care partners. This research would not have been possible without them. Finally, we acknowledge the research nurses who provided treatments throughout the trial.

## Disclosures

KEH is a founder of Resonance Therapeutics. PBF has received equipment for research from MagVenture A/S, Nexstim, Neuronetics and Brainsway Ltd and funding for research from Neuronetics. PBF is a founder of TMS Clinics Australia and Resonance Therapeutics. MRLE, NWB, CR, HC and FS have no biomedical financial interests or potential conflicts of interest to report.

---

**Supplementary Figure One.**
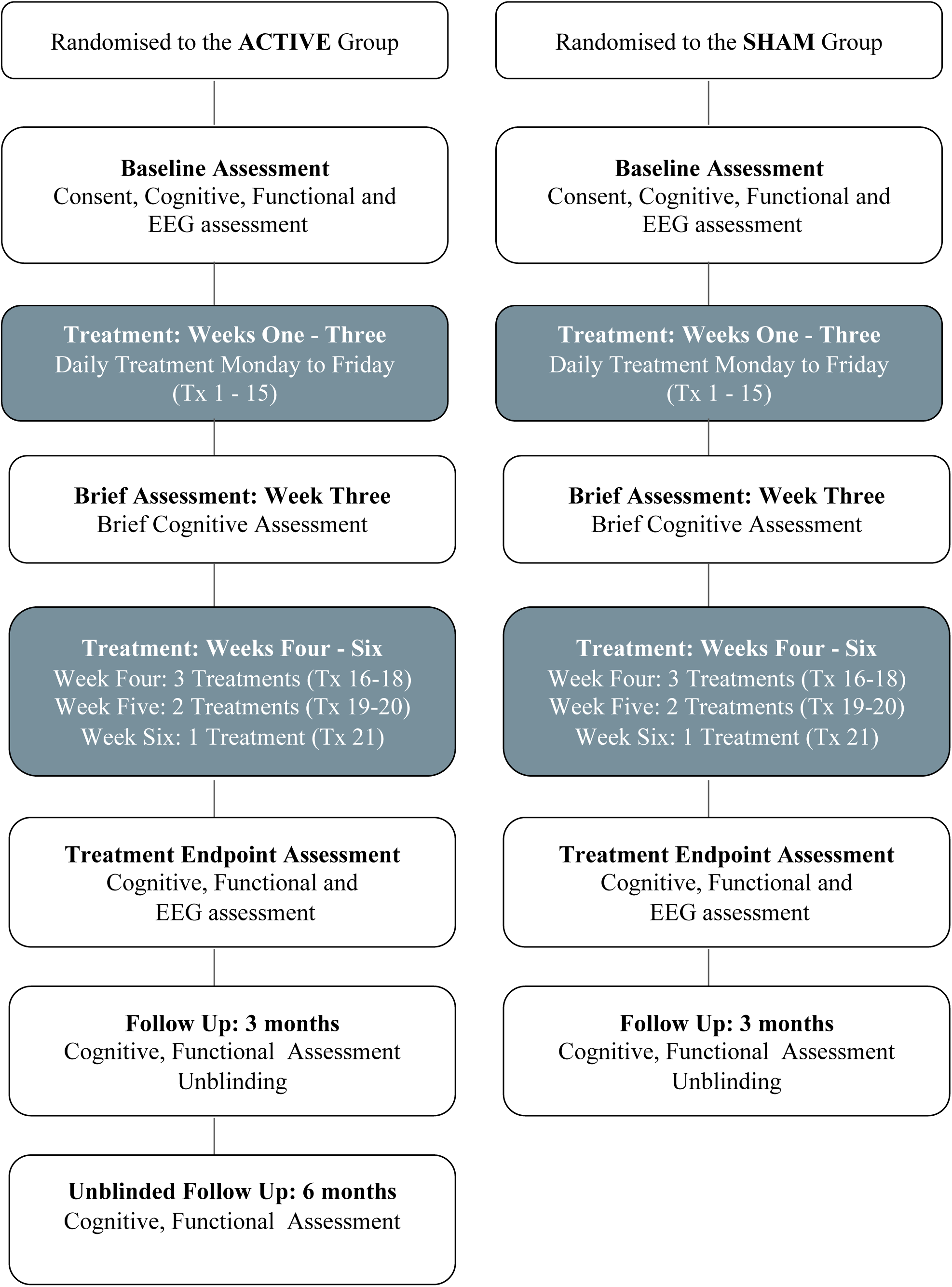

## Notes

### Clinical Trial

ACTRN12615000992505

### Author Declarations

Ethics Committee of Alfred Health gave ethical approval for this work on 8 Sep 2015 (ref. 372/15). Ethics Committee of Monash University gave ethical approval for this work on 30 Nov 2015 (ref.2015-2741-2550).

