## Supplementary Materials for "Gamma connectivity predicts response to intermittent Theta Burst Stimulation in Alzheimer’s disease: A randomised controlled trial"

### Materials and Methods

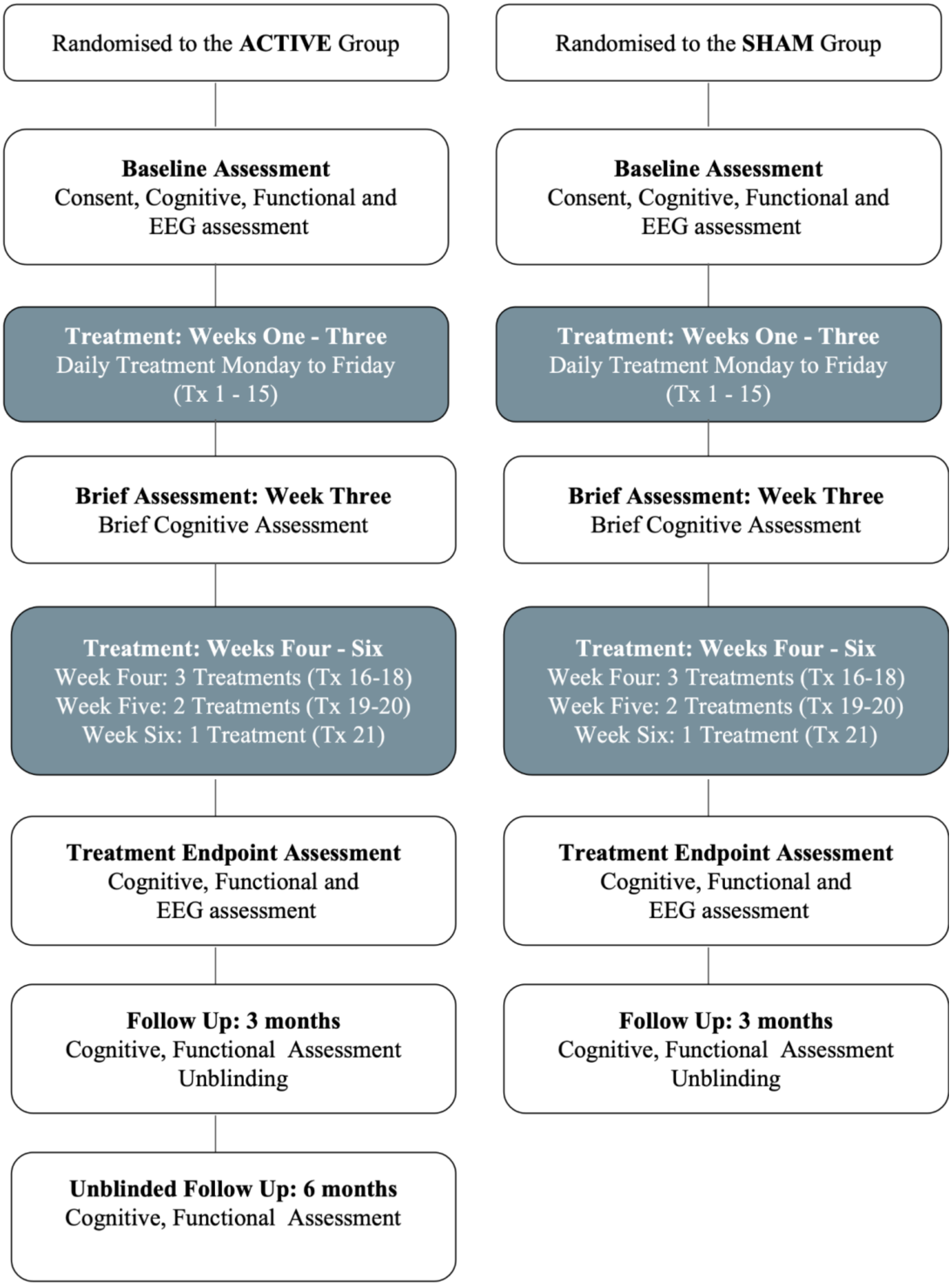


Supplementary Figure One. Study Flow Chart

#### ***Assessments***

*Resting state-EEG*

Specific electrodes used were AF3, AF4, F7, F5, F3, F1, FZ, F2, F4, F6, F8, FC5, FC3, FC1, FCZ, FC2, FC4, FC6, C5, C3, C1, CZ, C2, C4, C6, P7, P5, P3, P1, PZ, P2, P4, P6, P8, PO3, POZ, PO4, O1, OZ, O2, and M1, M2, SO1 (Reference: CPZ; Ground: AFZ).

Offline preprocessing was performed in MATLAB utilizing a fully automated cleaning pipeline, which we have recently validated against existing state-of-the-art EEG cleaning approaches and have determined that our pipeline cleans the artifacts more effectively while also preserving more of the brain activity signal. The pipeline is now in preparation for publication (Bailey et al., in preparation). The cleaned data is submitted to an independent component analysis, and artifactual components detected by IC Label are cleaned by wavelet enhanced ICA

#### ***Data Analysis***

*Resting state-EEG Functional Connectivity*

Data were epoched into 6 second segments (optimal for connectivity analysis, Miljevic et al. [2021]) with 2.5 second overlaps and Fourier transformed. The weighted Phase Lag Index (wPLI) computed in Fieldtrip (Oostenveld et al., 2011; Vinck et al., 2011) was used to examine resting state functional connectivity. The method used in the current paper adhered to previously published methods for extracting wPLI values (Bailey et al., 2018). First, instantaneous phase values were extracted from the complex Fourier spectra between 0.5 - 45 Hz by performing single Hanning taper time frequency transforms with a 1Hz resolution across a sliding time window corresponding to 7 oscillation cycles in length. These data were zero padded to ensure appropriate capture of the frequencies of interest.

Within each participant's data, and across all epochs, the instantaneous wPLI value was computed for each electrode pair using the debiased estimator method (Oostenveld et al., 2011). This measure of phase synchronisation weights connectivity computations against near-zero phase lag, and as such is robust against spurious connectivity from volume conduction of non-brain related noise artifacts and common reference artifacts (Bailey et al., 2018). Further, wPLI network characterization has been shown to exhibit good to excellent test-retest reliability (Hardmeier et al., 2014). Instantaneous wPLI values were derived at each measured time point and for each measured frequency of interest (0.5-45Hz), where wPLI values are bounded between 0-1 (0 indicating zero phase synchronisation; 1 indicating perfect phase synchronisation). It is important to note that due to the linear subtraction which occurs as part of the debiasing procedure it is possible to get negative values with wPLI. Within each participant, wPLI values were averaged across time within the middle 1 second of each epoch (which was unique to each epoch given the epoch overlap settings), and then averaged within the two frequency bands of interest (theta, 4-8Hz, gamma 30-45Hz). Between-group differences in frequency band specific functional connectivity were then statistically analysed using non-parametric cluster-based permutation statistics implemented by the Network Based Statistic (NBS) (Zalesky et al., 2010).

### Results

#### *Resting State-EEG: Functional Connectivity*

Supplementary Table One. Means and standard deviations of averaged gamma wPLI values from the identified network across both treatment groups at each timepoint.

| | | **Baseline** | | | **End of Treatment** | | |
|---|---|---|---|---|---|---|---|
| | | **n** | **Mean** | **sd** | **n** | **Mean** | **sd** |
| **Gamma wPLI** | **Active** | 25 | 0.0025 | 0.0145 | 25 | 0.0147 | 0.0187 |
| | **Sham** | 23 | 0.0025 | 0.0075 | 23 | -0.0010 | 0.0103 |

#### *Cognitive and Functional Outcomes*

Supplementary Table Two. Means and standard deviations of cognitive and functional assessments across both treatment groups at each timepoint.

| | | Baseline | | | Week 3 | | | End of Treatment | | | 3 month follow up | | | 6 month follow up | | |
|---|---|---|---|---|---|---|---|---|---|---|---|---|---|---|---|---|
| | | **n** | **Mean** | **sd** | **n** | **Mean** | **sd** | **n** | **Mean** | **sd** | **n** | **Mean** | **sd** | **n** | **Mean** | **sd** |
| **ISL Delayed Recall** | **Active** | 28 | 0.96 | 1.42 | 26 | 1.30 | 1.64 | 23 | 1.95 | 2.14 | 19 | 1.63 | 1.38 | 11 | 2.00 | 2.00 |
| | **Sham** | 28 | 1.10 | 1.03 | 26 | 2.19 | 1.95 | 26 | 0.96 | 1.28 | 21 | 1.33 | 1.77 | na | na | na |
| **ISL Total** | **Active** | 28 | 10.32 | 4.48 | 26 | 11.07 | 4.27 | 23 | 11.86 | 4.94 | 19 | 10.00 | 3.34 | 17 | 13.70 | 5.04 |
| | **Sham** | 28 | 9.25 | 5.56 | 26 | 11.61 | 7.16 | 26 | 11.03 | 6.89 | 21 | 10.85 | 4.73 | na | na | na |
| **ADASCog** | **Active** | 28 | 20.79 | 9.17 | na | na | na | 23 | 20.20 | 10.26 | 19 | 20.47 | 9.41 | 19 | 16.04 | 8.82 |
| | **Sham** | 28 | 22.16 | 12.69 | na | na | na | 27 | 22.29 | 12.55 | 23 | 21.93 | 12.86 | na | na | na |
| **QoL-AD (self)** | **Active** | 28 | 41.50 | 4.49 | na | na | na | 23 | 42.48 | 5.350 | 19 | 40.11 | 5.07 | 19 | 41.05 | 6.13 |
| | **Sham** | 28 | 38.79 | 6.22 | na | na | na | 27 | 39.96 | 6.309 | 23 | 36.48 | 7.03 | na | na | na |
| **QoL-AD (carer)** | **Active** | 26 | 37.23 | 6.44 | na | na | na | 12 | 38.67 | 5.24 | 13 | 37.08 | 3.73 | 10 | 34.70 | 5.92 |
| | **Sham** | 23 | 35.17 | 5.89 | na | na | na | 14 | 35.14 | 4.03 | 11 | 31.55 | 4.50 | na | na | na |
| **GDS** | **Active** | 28 | 2.25 | 1.57 | na | na | na | 21 | 2.14 | 1.90 | 19 | 2.32 | 2.49 | 19 | 2.00 | 2.05 |
| | **Sham** | 28 | 2.46 | 1.89 | na | na | na | 27 | 2.41 | 1.94 | 21 | 1.81 | 1.63 | na | na | na |

ISL: International Shopping List; ADASCog: Alzheimer's Disease Assessment Scale – Cognition; QoL-AD: Alzheimer's Disease Quality of Life; GDS: Geriatric Depression Scale.

#### *Follow up cognitive and functional outcomes*

*3-month follow-up*

For the 3-month follow up data, LMMs were conducted for the cognitive and functional variables (i.e. ISL Delayed Recall, ISL Total, ADASCog, QoL-AD [self, carer], GDS), with fixed effects of group (active, sham) and time (end of treatment, three month follow up). There were no significant main or interaction effects across the follow up period (end of treatment to 3 Month follow up) for ISL Delayed Recall, ISL Total, ADASCog, QoL-AD (carer) and GDS. See Supplementary Table Three. For the ISI Delayed Recall this would indicate that the improvement seen from Baseline to End of Treatment was not significantly reduced by the 3 month follow up.

For the QoL-AD (self) there was a significant main effect of Time ($F_{(1, 42.114)}$ =6.814, p = 0.012) and no interaction effect ($F_{(1, 42.114)}$ =0.314, p = 0.578). Post-hoc analyses revealed a decrease in QoL-AD over time, irrespective of treatment group, from End of Treatment to 3 month follow up (mean difference = -1.911, p = 0.012). See Supplementary Table Three.

Supplementary Table Three. Linear Mixed Models of Exploratory Outcomes in the Follow Up Phase (End of Treatment to 3 month follow up).

| **Assessment** | **Effect over Time** | **p value** | **Time by Group Effect** | **p value** |
|---|---|---|---|---|
| ISL-Delayed Recall | F (1, 40.448) = 0.382 | 0.540 | F (1, 40.448) = 0.655 | 0.423 |
| ISL-Total | F (1, 41.914) = 2.559 | 0.117 | F (1, 41.914) = 1.330 | 0.255 |
| ADASCog | F (1, 44.039) = 0.087 | 0.769 | F (1, 44.039) = 0.00 | 0.998 |
| QoL-AD self | F (1, 42.114) = 6.814 | 0.012* | F (1, 42.114) = 0.314 | 0.578 |
| QoL-AD carer | F (1, 21.616) = 1.172 | 0.291 | F (1, 21.616) =0.205 | 0.655 |
| GDS | F (1, 42.113) = 1.091 | 0.302 | F (1, 42.113) = 1.303 | 0.260 |

ISL: International Shopping List; ADASCog: Alzheimer's Disease Assessment Scale – Cognition; QoL-AD: Alzheimer's Disease Quality of Life; GDS: Geriatric Depression Scale.

*6 Month follow up (active group only)*

For the 6 month follow up data, collected for participants in the active group only, we again conducted LMMs to investigate any changes from 3 to 6 month follow up. From the 21 participants in the active group who completed the 3 month follow up 19 returned for the 6 month follow up. There were no significant changes from 3 to 6 months on any of the cognitive and functional assessments with the expectation of the ISL-Total, which showed a significant increase from 3 to 6 months. see Supplementary Table Four

Supplementary Table Four. Linear Mixed Models of Exploratory Outcomes in the Follow Up Phase, active group only (3 to 6 month follow up).

| **Assessment** | **Effect over Time** | **p value** |
|---|---|---|
| ISL-Delayed Recall | F (1, 11.741) = 0.006 | 0.939 |
| ISL-Total | F (1, 17.198) = 6.965 | 0.017* |
| ADASCog | F (1, 18.682) = 1.443 | 0.245 |
| QoL-AD self | F (1, 19.944) = 0.241 | 0.629 |
| QoL-AD carer | F (1, 10.969) = 0.876 | 0.370 |
| GDS | F (1, 19.966) = 0.166 | 0.668 |

ISL: International Shopping List; ADASCog: Alzheimer's Disease Assessment Scale – Cognition; QoL-AD: Alzheimer's Disease Quality of Life; GDS: Geriatric Depression ScalE
